# Risk of COVID-19 Reinfection and Vaccine Breakthrough Infection, Madera County, California, 2021

**DOI:** 10.1101/2022.01.22.22269105

**Authors:** Minhphuong Nguyen, Eric Paul, Paul K. Mills, Simon Paul

## Abstract

The probability of either testing COVID-19 positive or dying for three cohorts in Madera County, California in 2021 was compared. These cohorts included 1. those unvaccinated, 2. those vaccinated and 3. persons with a previous COVID-19 infection. The three groups were made generally comparable by matching on age, gender, postal zip code of residence, and the date of either COVID-19 infection or of vaccination.

The hazard ratio (HR) for death (from all causes) after COVID-19 infection vs. vaccination was 11.7 (95% CI 5.91-23.1, p<0.05). The HR for testing positive for COVID-19 >14 days after initial COVID-19 infection or after completing primary COVID-19 vaccination was 1.98 (95% CI 1.53-2.58 p<0.001). As the majority of positive COVID-19 tests in the post COVID-19 cohort occurred within 90 days of the initial infection, and as these early positives may not represent a new infection, we also compared rates of testing COVID-19 positive ≥ 90 days after initial infection or vaccination. After removing these early positive COVID-19 tests that occurred between days 14-90, the HR ratio for testing COVID-19 positive is now lower for the post COVID-19 cohort compared with the vaccinated cohort. The risk for having a positive COVID-19 test occurring 90 days after an initial COVID-19 infection or after vaccination was 0.54 (95% CI 0.33-0.87, p<0.05) for the post COVID-19 group vs Vaccinated group.

Thus the risk for testing COVID-19 positive was higher in the first 90 days after COVID-19 infection compared to those vaccinated. However, from 90 to 300 days after COVID-19 infection, the post COVID-19 infection cohort had a lower risk of testing COVID-19 positive than those fully vaccinated.

## Introduction

Madera County is located in the agricultural Central Valley region of California, population 158,217. The County is majority White-Hispanic (58.5%) and has had a fairly high incidence of COVID-19 during 2021 compared with other areas of California (23.6 cases/100,000 person-days on average for the time period of this study). The level of vaccination county-wide has reached only 50% as of January, 2022, and as of this date 16% of the county has had a documented COVID-19 infection.(*1*) This relatively high rate of ongoing COVID-19 infections and moderate level of vaccination makes Madera County a promising site for comparing the community rates of both COVID-19 reinfection and vaccine breakthrough infections.

Determining the risk of reinfection after a primary infection with COVID-19 is important for risk assessment, modeling of future COVID-19 incidence, for prioritization of vaccination education and resources, and for the timing of required compliance post COVID-19 infection with vaccination mandates. Immunologic studies provide results more rapidly than studying clinical outcomes, however, while studies such as those of antibody titers after infection or vaccination may indicate different levels of immunity and changes in immunity, there are not well established clinical correlations for these in-vitro results and disease.(*2*) For clinical studies of reinfection rates and vaccine breakthrough rates there are other significant obstacles in addition to the time required for clinical data to accrue. Prospective studies of specific groups such as health care workers can provide high quality data but may be limited in generalizability.(*3*–*5*) Community level observational studies may be biased as they are based on subject-driven testing. Persons with varying degrees of access to and motivation for testing for COVID-19 when symptomatic will differ. There are also unknown levels of undiagnosed prior COVID-19 in the community, and changing levels of COVID-19 incidence and COVID-19 variants in the community over time.(*3*)

The primary goal of this study was to compare the risk of re-infection in persons recovered from an initial COVID-19 infection to the risk for persons who were fully vaccinated. The benefit of full vaccination has been well studied in randomized clinical trials (i.e. vaccine efficacy) and also in large observational studies (i.e. effectiveness),(*6*) so the protection from full vaccination can serve as a benchmark for comparison. To address various critical real-world issues of changing COVID-19 incidence over time, new variants emerging over time, and the difference in access and willingness to test in different populations, we chose to study the incidence of COVID-19 infections in three matched cohorts. Initially we selected three groups of persons from the available testing and vaccination data in Madera County. These groups were the “unvaccinated” group consisting of persons who have tested COVID-19 negative prior to the start of the study (January 1, 2021) and who were not vaccinated. The criterion of using a prior negative test was partly for convenience as the testing registration process elicited demographic information for this population. This information is not available for the untested, unvaccinated population at large in the County. By testing negative, persons in this group have demonstrated access to testing, and possibly a level of risk and concern for COVID-19 higher than that of the never-tested population. The second group, the “Vaccinated,” consists of persons who had either never been tested for COVID-19 or had tested negative for COVID-19 prior to vaccination. The “post COVID-19” group includes all people who tested positive for the first time on or after January 1, 2021.

There were significant differences in the characteristics of these three groups, most importantly age (due to the age-based rollout of vaccine eligibility) and also the distribution of dates of testing positive for COVID-19 in 2021 (as there were few vaccinated persons in the first 4 months of 2021). As there could be significant bias from comparing mortality rates in groups of differing ages, and infection rates if different time periods were compared, three matched cohorts were selected from each of the three groups. These three cohorts were created by selecting persons from each of the three groups that had identical gender, postal zip code of residence, age (+/- 2 years) and date of infection or completing vaccination (+/- 10 days). Residence postal code was included as a matching criterion as a proxy for socioeconomic status and race. Data on socioeconomic status was not available for individuals and race data from vaccine and test registration is often incomplete (especially for earlier time periods) which would make matching difficult. However, as these demographic variables for Madera County population are correlated with location of residence, we used matching by postal code in an effort to match for race and socioeconomic status. The goal of matching people for the final criterion, date of vaccination or of COVID-19 infection, was to ensure that identical time periods of risk were being observed for each cohort.

## Methods

COVID-19 testing results (negative and positive, including antigen tests, molecular testing and COVID-19 antibody tests) must be reported to the California Reportable Disease Exchange system (CalREDIE).(*7*) Commercial, public health and most clinical laboratories report results electronically directly to CalREDIE. Rapid antigen testing may be reported by manual entry through internet or app-based access. The extent to which home self-testing results are reported is not known. All reported COVID-19 test results with demographic information in CalREDIE for residents of Madera County, California were accessed. COVID-19 Immunization data for California are reported to a statewide database SNOWFLAKE.(*8*) Death data for Madera County are reported directly to the Department of Public Health Vital Statistics department.

Persons were identified and all duplicates were removed by grouping results based on a unique DOB, gender and first three initials of their first and last names. For each person the following information was determined: their first and last COVID-19 positive test dates, their first COVID-19 negative test date, their vaccination status and administration dates, whether they completed the primary vaccine series, and their date of death if deceased (all cause mortality). Vaccine recipients who had no COVID-19 test data were also included in this dataset.

Thus, these are the selection criteria for the three subject groups.

Group 1--” Unvaccinated, with prior COVID-19 test negative”

Persons who had received a negative COVID-19 test result prior to 1/1/2021, who had no positive COVID-19 test results prior to 1/1/2021, and who had not received any vaccination doses during the study time period.

Group 2--” Vaccinated”

Persons who had completed a primary COVID-19 vaccination series on or after 1/1/2021 and who had either no COVID-19 test results or only negative test results prior to 1/1/2021.

Group 3: “Post COVID-19 Infection”

Persons who had either no COVID-19 test results or only negative COVID-19 test results prior to a documented positive COVID-19 antigen or molecular test that was positive on or after January 1, 2021, and who had received no doses of COVID-19 vaccine during the study time period.

Any positive COVID-19 antibody test result prior to the first positive COVID-19 test date was also an exclusion criterion for all three groups.

### Matching

There were significant differences in the demographic characteristics of these three groups, most importantly age (due to the age-based rollout of vaccine eligibility) and distribution of dates of testing positive for COVID-19 in 2021 (as there were few vaccinated persons in the first 4 months of 2021). As there could be significant bias from comparing mortality rates in groups of differing ages, and infection rates if different time periods were compared, three matched cohorts were selected from the three groups. These three cohorts were created by selecting persons from each of the three groups that had identical gender, postal zip code of residence, age (+/- 2 years) and date of infection or completing vaccination (+/- 10 days). Residence postal code was included as a matching criterion as a proxy for socioeconomic status and race. Using test and vaccine registration information, data on socioeconomic status was not available for individuals and race data is often incomplete (especially for earlier time periods) which would make matching difficult. However, as differences in these demographic variables for Madera County population are correlated with location of residence, we used matching by postal code in an effort to match for race and socioeconomic status. The goal of selecting matching persons based on date of vaccination or of COVID-19 infection was to ensure that identical time periods of risk were being observed for each cohort. Of note, of the 9,571 persons in the post COVID-19 infection group, 3253 did not have a match in the vaccinated group. This inability to find vaccinated matches is expected for younger COVID-19 cases diagnosed in the early months of 2021: these cases would not have a vaccinated match as those younger age groups did not become eligible for vaccination until later in the year.

Persons in Groups 2 and 3 were matched as follows. One person was selected at random from Group 2, and then the first Group 3 person that matched this Group 2 person on DOB (+/- 730 days), gender, postal zip code, and first positive COVID-19 test collection date (+/- 10 days of Vaccine series completion date) was randomly selected. These persons were then removed from the groups and the matching process repeated until all possible matches meeting these four criteria were obtained. Persons in Group 1 do not have a COVID-19 positive test date or vaccination date to use for matching. Therefore, members of Group 1 were selected at random after meeting the same matching criteria as the Group 2&3 subject pairs already selected (i.e. the group 1 candidate matched on the three criteria of the Group 2 person: DOB +/- 730 days, sex, postal zip code). The Group 1 person would be rejected if they had a death or positive COVID-19 test date prior to the date used for matching groups 2 and 3 (COVID-19 positive test date or vaccination completion date).

### Outcome Events (infection and/or death)

Cohort 1 unvaccinated prior COVID-19 test negative: the date of group 2 vaccination that was used for matching cohorts 2 and 3 was also used as the initial date to begin recording events in cohort 1. This date was chosen as the start date for cohort 1 for recording events to ensure that identical time periods at risk were used for observing events in the three groups. Deaths were recorded on or after this matching date. Infection events were recorded starting after day 14 (or after day 90) from this matching date. These recording start dates were chosen to be identical with the event recording time-period of cohorts 2 and 3.

Cohort 2 vaccinated: Death events were recorded beginning on or after the day of the final dose administration for the person’ s primary vaccination series. Infection events were recorded after 14 days (or 90 days) post completion of the vaccination series, to allow patients to achieve full immunity post vaccination (14 days) and for comparison with the infection event recording of cohort 3 at 90 days consistent with the California Department of Public Health (CDPH) definition of reinfection post COVID-19.(*9*) If there were multiple positive test results after completing vaccination, the first positive test result date was selected as the event date.

Cohort 3 post COVID-19 infection: deaths events were recorded beginning on the date of collection of the person’ s first positive COVID-19 test result. Infection events were recorded beginning 14 days after the initial positive test collection date (to allow for the subject to complete isolation from their COVID-19 infection, to develop immunity post COVID-19 and to match the 14-day time period allowed for developing full immunity in the vaccinated group). If multiple positive tests were recorded after day 14, the last positive test date was considered the event date for cohort 3. Infection events occurring after 90 days were also evaluated to match the CDPH definition of reinfection.

COVID-19 testing in Madera County 2021 was largely personal choice for the general population (based on symptoms or concern about exposure). Some surveillance testing was required of unvaccinated healthcare personnel (those with exceptions to mandated vaccination) which also contributed testing results. For certain high risk youth sports weekly surveillance testing was carried out from April 6-June 14 2021.(*10*) There was no school surveillance testing in Madera County in 2021, however, beginning in September 2021 a significant volume of testing came from school contact tracing. After August of 2021 directives from the California Department of Health mandated COVID-19 testing for specific job occupations, visitors to health care facilities, for unvaccinated workers in healthcare and for staff in school environments.(*11*–*13*) These mandated test results are included in the CalREDIE database and were included in our study dataset. There was no COVID-19 testing carried out as a part of this study.

Vaccination in Madera County during 2021 was initially restricted based on age, health status, and occupation. By May 12, 2021, vaccination was available to all those ≥ 16 years old. Ages 5-11 were not eligible for vaccination prior to the end date of this study (12/7/2021).(*14*)

Risk of “failure” in the three above defined risk groups was evaluated using a survival analysis approach. “Failures,” including infection and death, were recorded beginning at the time points defined above. The Kaplan Meier survival function was calculated as events occurred (deaths for the mortality KM (figure 2), and infections for the testing positive KM curves (figures 3-5). Survival curves were plotted for each group.(*15*) Data were censored for death or reaching the study end-date (12/7/2021) for the mortality hazard curve. Data for the testing COVID-19 positive curves were censored for death or reaching the study end-date (12/7/2021).(*15*) Differences in hazard ratios between cohorts were statistically tested using the log rank test.(*16*) Survival analyses were computed using lifelines survival analysis library(*17*) and plotted using the matplotlib library for python 3.9.(*18*)

**Figure 1:**
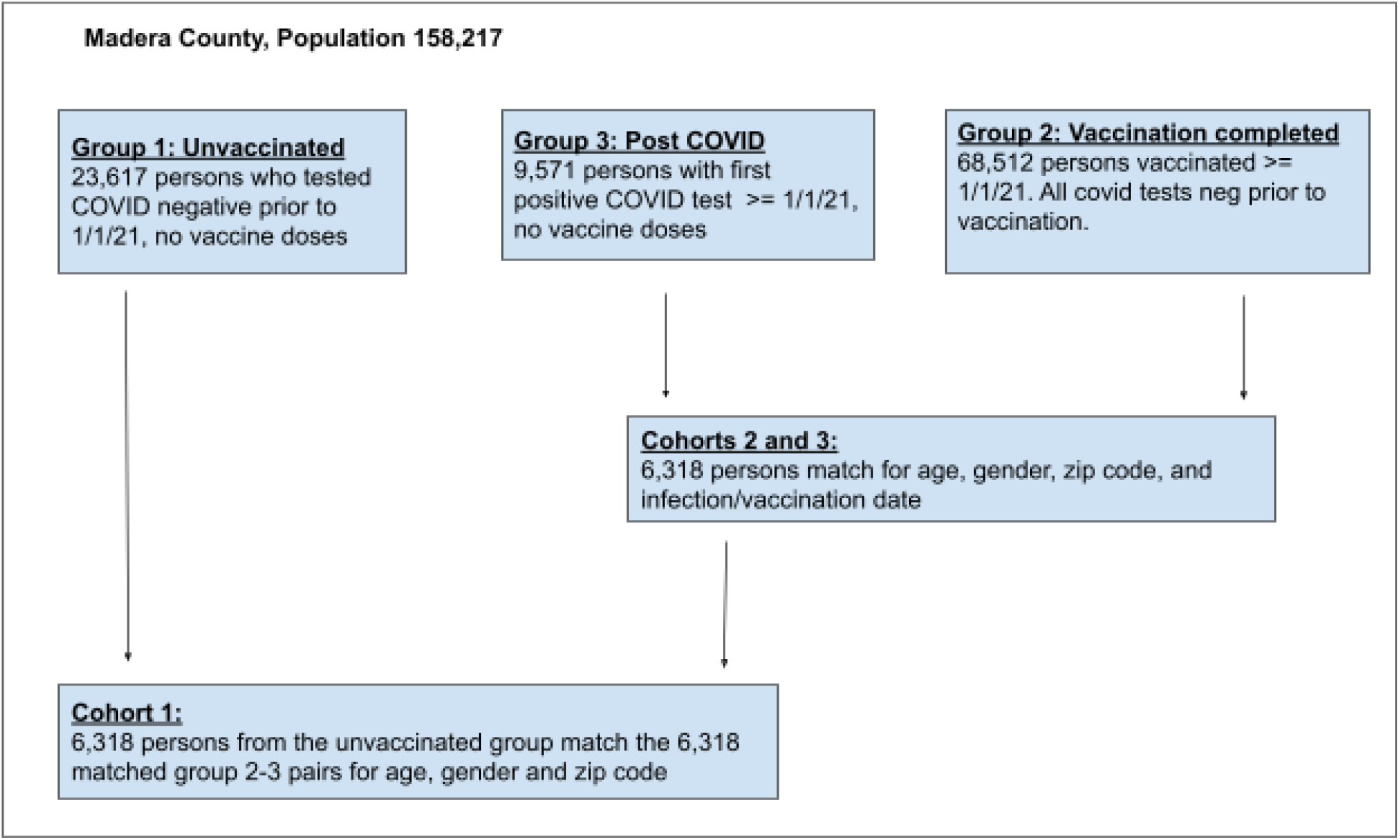
Schema of selection and matching process

**FIgure 2:**
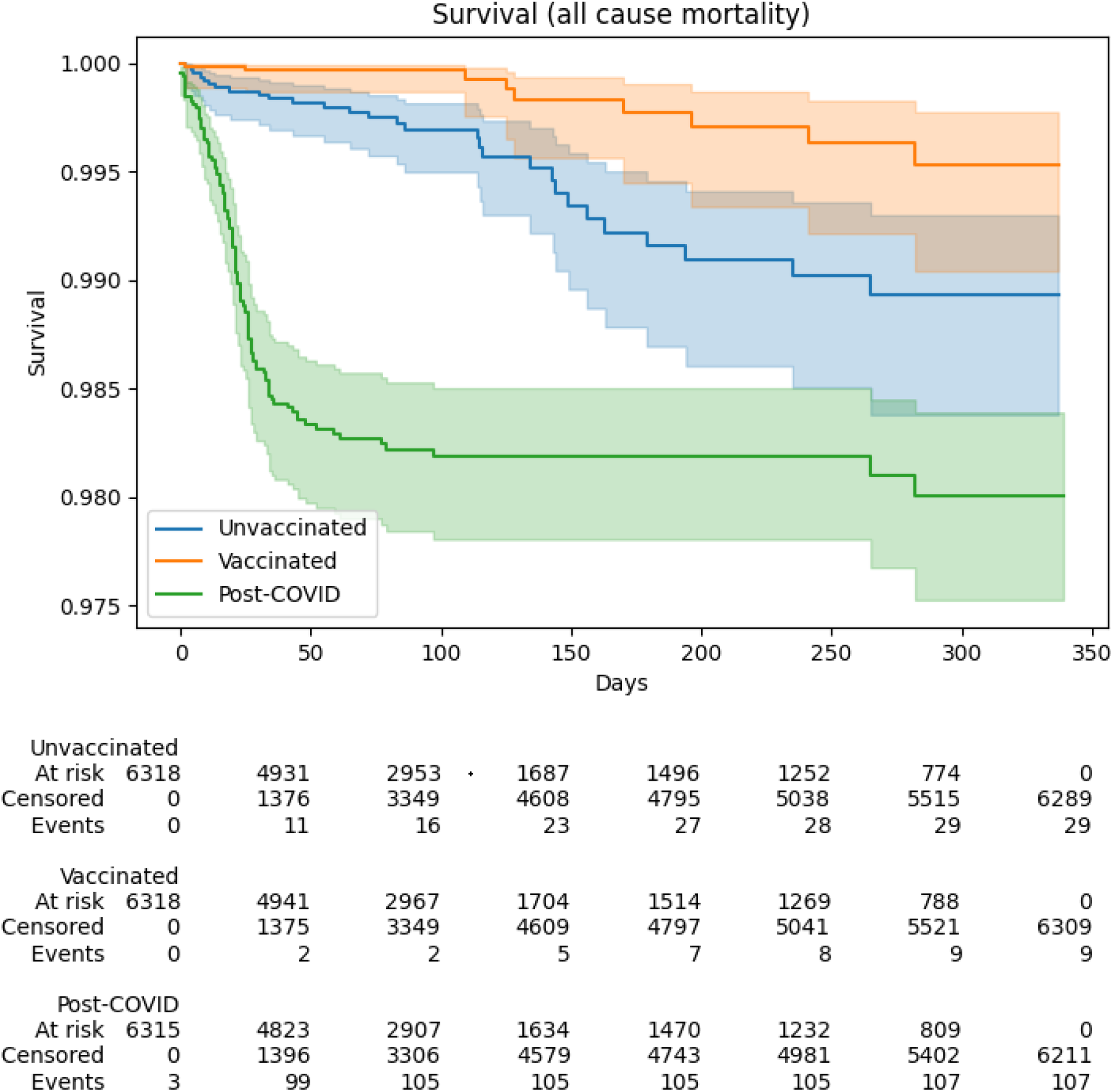
Probability of Survival over time for the three Cohorts

**Figure 3:**
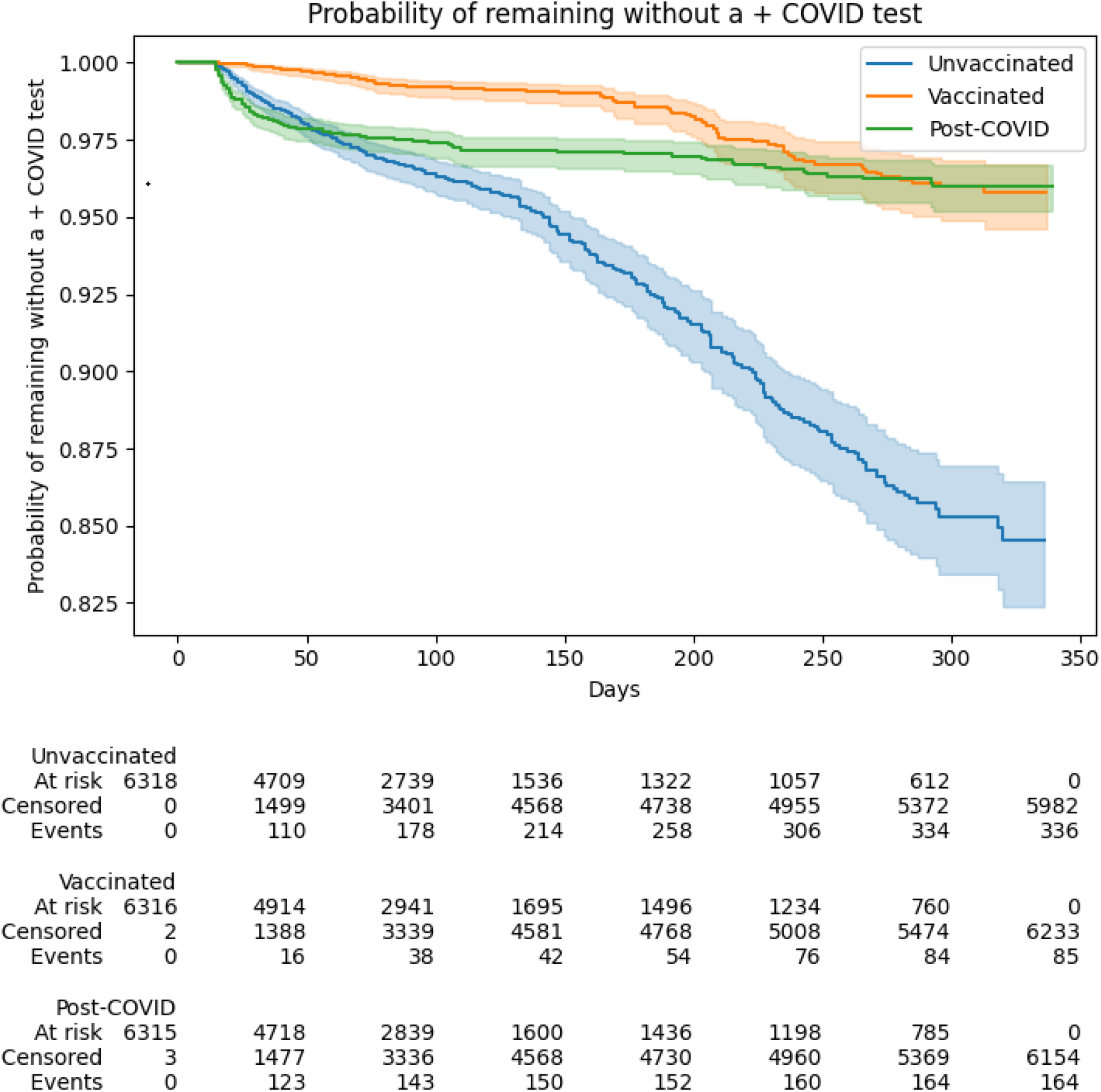
Probability of remaining without a positive COVID-19 test over time for the three Cohorts, beginning 14 days after initial infection or vaccination

To compare risk of failure in the unvaccinated and post COVID-19 infected groups to the vaccinated cohort, hazard ratios (HR) and 95% confidence limits were calculated using Stata software.(*16*)

Chi-Square calculations were performed using web based calculators.(*19*)

## Results

Testing volume varied significantly over time (testing increased during times of increased COVID-19 incidence) and also increased with school reopening in the Fall of 2021. During the time period of this study, the average rate of COVID-19 testing in Madera County was 383 tests/100,000 person-days. There were no reported cases of the omicron variant in Madera County during the time period of this study.

### Matching of the three cohorts

Prior to matching, the average age of the vaccinated group was significantly older (48 years) compared with the unvaccinated and post COVID-19 groups (37 and 31 years respectively) (see Table 1).

**Table 1:** Demographics of Madera County, the selected Unvaccinated, Vaccinated and Post COVID-19 groups, and the matched cohorts drawn from these three groups

|  |  | Selected Groups: |  |  | Matched Cohorts: |  |  |
| --- | --- | --- | --- | --- | --- | --- | --- |
|  | Madera Co. | Unvaccinated | Vaccinated | Post-COVID | Unvaccinated | Vaccinated | Post-COVID |
| n | 158217 | 23617 | 68512 | 9571 | 6318 | 6318 | 6318 |
| % Female | 51.8% | 54% | 53% | 51% | 52% | 52% | 52% |
| Age (mean) | 34.1 | 37 | 48 | 31 | 35.5 | 35.5 | 35.4 |
| Race/Ethnicity |  |  |  |  |  |  |  |
| Hispanic<sup>1</sup> | 58.8% |  |  |  | 20.1% | 54.3% | 51.3% |
| White | 33.2% |  |  |  | 17.9% | 28.3% | 30.3% |
| African American | 4.2% |  |  |  | 1.1% | 2.2% | 2.8% |
| Other | 3.8% |  |  |  | 2.0% | 6.0% | 3.7% |
| Unknown | n/a |  |  |  | 59.0% | 9.2% | 11.9% |

<sup>1</sup>Only White-Hispanics were reported in the three matched cohorts

The matching process identified 6,318 persons in each group who had a matching age, gender, postal code and date of infection/vaccination (if more than one person could be matched, then a single match was selected at random). From the 9,571 people who tested positive for COVID-19 in 2021 there were 6,318 having a corresponding person in the vaccinated group who: was vaccinated the same week as the week a person tested positive, and who was of the same age, gender and lived in the same postal code as the person who tested positive. That is, of the 9,571 post COVID-19 persons and the 68,512 vaccinated persons, there were 6,318 persons who had a matching age, gender, zip code AND week of COVID-19 infection or week of vaccination. For each of these 6,318 matched pairs of post COVID-19 to vaccinated persons, a matching person of the same age, gender and postal code of residence was present within the 23,617 persons in the unvaccinated Group 1.

The average age of these groups was 36 years old, and 52% were female (see Table 1). Race and ethnicity were not used for matching as there were many persons missing this demographic information. However, a comparison of the available demographics for the three cohorts after matching using the available race and ethnicity data is possible. As Cohort 1 (unvaccinated) included all persons with negative COVID-19 tests results beginning in 2020, and 2020 was when race and ethnicity data was most often not recorded, Cohort 1 has by far the largest percent of subjects missing race/ethnicity data (59% missing data for Cohort 1, vs 9,2% and 11.9% for Cohorts 2 and 3). The data in Table 1 demonstrates how the use of zip code for matching resulted in similar race/ethnicity composition for Cohorts 2 and 3. Cohort 3 has just 2% more non-Hispanic white subjects, and 2.7% more of unknown ethnicity (the differences in race/ethnicity data between groups 2 and 3 is statistically significant, however, p<0.01 by chi-square analysis).(*19*)

The demographics of Madera County overall, and the three groups prior to and after matching are shown in Table 1.

### Death and COVID-19 testing results

The number of deaths and positive tests for Madera County as a whole and the three matched cohorts are shown in Table 2.

**Table 2:** Outcomes for Madera County, the three selected groups, and the matched cohorts from each of these three groups.

|  | Madera Co. * | Selected Groups: |  |  | Matched Cohorts: |  |  |
| --- | --- | --- | --- | --- | --- | --- | --- |
|  |  | Unvaccinated | Vaccinated | Post-COVID | Unvaccinated | Vaccinated | Post-COVID |
| Deaths (all cause) | 3018 | 206 | 329 | 164 | 29 | 9 | 107 |
| COVID+ test p 14 Days | 13577 | 1638 | 1584 | 250 | 336 | 85 | 164 |
| COVID+ test p 90 Days |  | 1199 | 1405 | 56 | 169 | 48 | 25 |
| Deaths/100K person-days | 5.2 | 2.7 | 2.4 | 11.7 | 3.8 | 1.1 | 13.6 |
| COVID+ post 14 Days/100K person-days | 23.6 | 21.7** | 11.3 | 17.8 | 44.6** | 10.5 | 20.8 |
| COVID + post 90 Days/100K person-days |  | 15.9 | 10.0 | 4.0 | 22.4 | 5.9 | 3.2 |

\*for Madera County data no 14 day lag was included: 13,577 is the total number of all COVID-19 positive tests for 1/1/2021-12/7/2021 and the incidence rate of 23.6/100,00 person days is based on that number of infections.
\*\* for the unvaccinated group prior to matching, a start date for observation of 1/1/2021 was used. After matching and assigning a start date matched to the vaccination date for the matched pair, the mean start date for the unvaccinated cohort changed to 7/7/2021. This shift in mean start date moved the observation period which then corresponded with the fall surge in infections, thus resulting in a markedly higher incidence rate post matching.

The initial COVID-19 infection that led to inclusion in the post COVID-19 cohort had 99 deaths in the first 50 days (See Figure 2 survival KM)--markedly higher than in the vaccinated and unvaccinated cohorts. The hazard ratio (HR) for death (all causes) in the post COVID-19 infection cohort compared to the vaccination cohort was 11.7 (95% CI 5.91-23.1, p<0.05). However, for those that survived the initial COVID-19 infection and remained in the post COVID-19 cohort, significant protection from retesting COVID-19 positive was then demonstrated (see Figures 3,4). This protection was observed especially after the initial 90-day period where PCR tests may remain positive without clinical reinfection. The probability of testing COVID-19 positive remained lowest for the post COVID-19 cohort for the time period from day 90 to day 300 (Figure 4).

**Figure 4:**
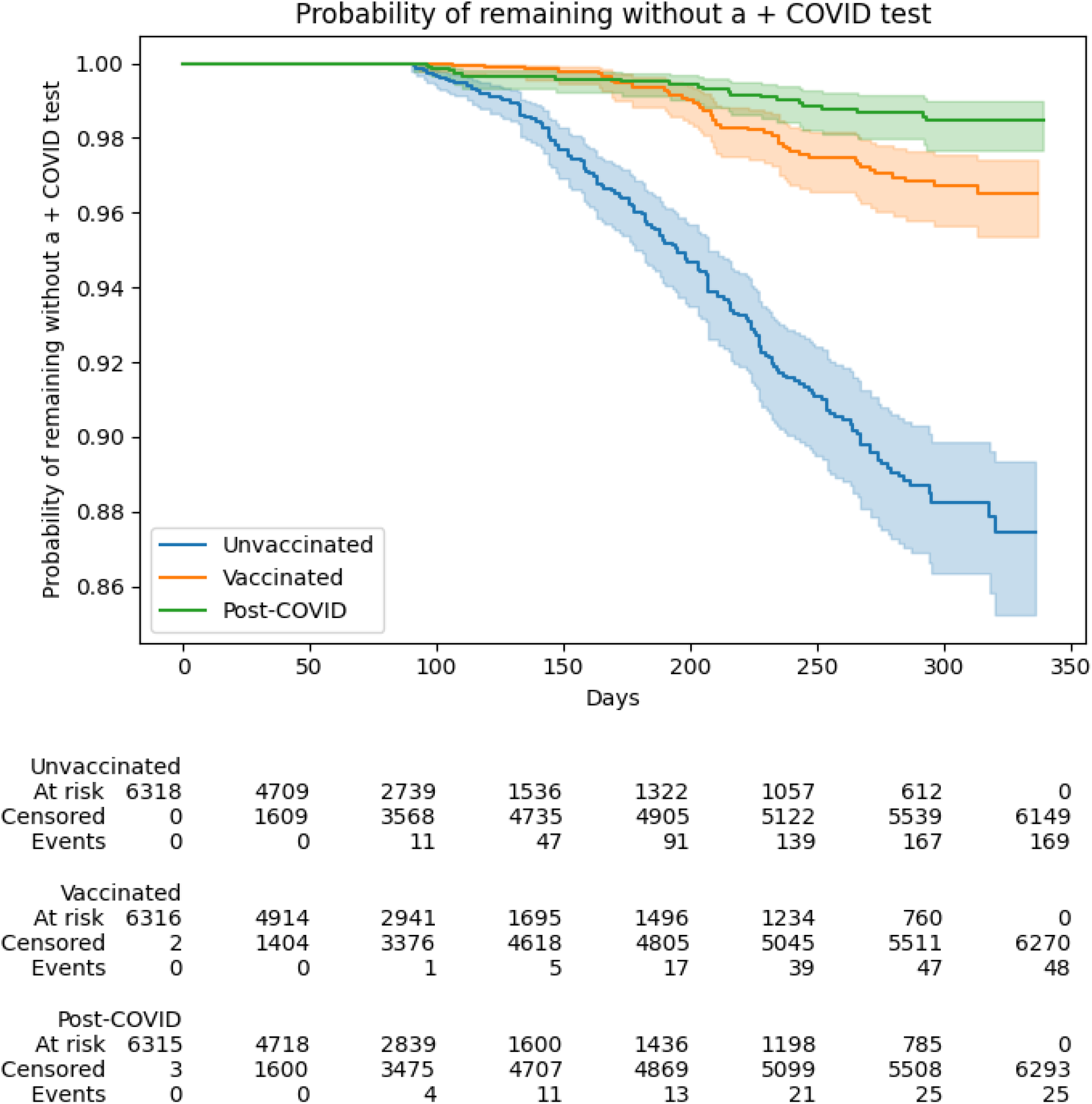
Probability of remaining without a positive COVID-19 test over time for the three Cohorts, beginning 90 days after initial infection or vaccination

The HR for testing positive for COVID-19 positive >14 days after initial COVID-19 infection compared with after completing primary COVID-19 vaccination was 1.98 (95% CI 1.53-2.58 p<0.001). As positive COVID-19 tests within 90 days of an initial infection may not represent a new infection, rates of testing COVID-19 positive ≥ 90 days after initial infection or vaccination were also compared. The HR for a positive COVID-19 test occurring 90 days post infection or vaccination was 0.54 (95% CI 0.33-0.87, p<0.05)--that is, a lower rate for testing positive in the post COVID-19 cohort than in the vaccinated cohort.

The high-risk subgroup of those ≥55 years old, who may be expected to mount a less vigorous immune response either to a prior infection or vaccination, was also evaluated. The risk of testing COVID-19 positive after 90 days post infection or vaccination for those ≥55 years old was similar to the probability for the entire cohorts including all ages. For those over age 55, there were the fewest positive COVID-19 tests after day 90 for the post COVID-19 cohort. However, with the now smaller cohort size (1,048 persons) this difference favoring the post COVID-19 cohort was not statistically significant (Figure 5).

**Figure 5:**
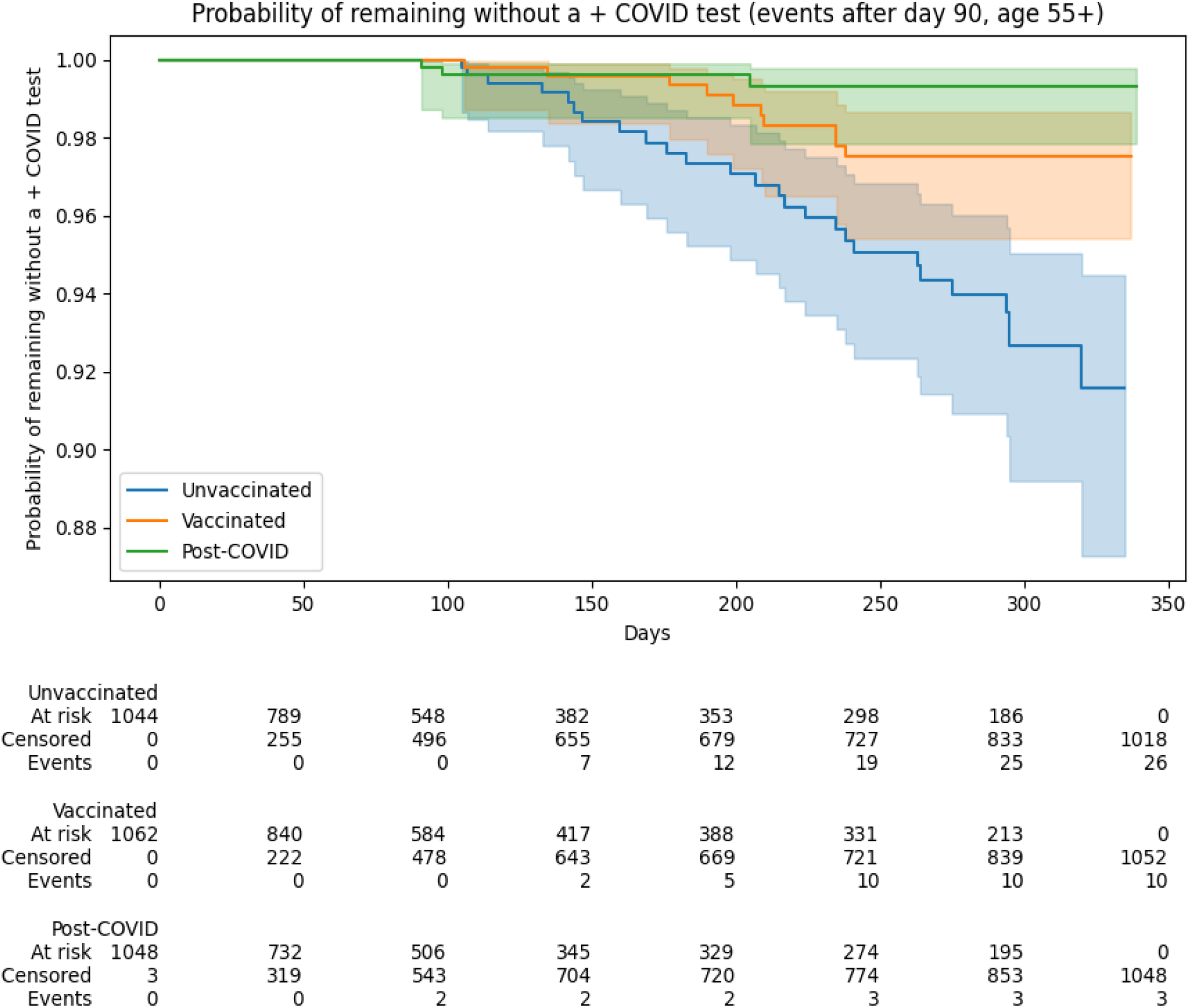
Probability of remaining without a positive COVID-19 test over time for persons ≥ 55 years-old in the three Cohorts, beginning 90 days after initial infection or vaccination

## Discussion

In this analysis, the risk of death after testing positive for COVID-19 was, as expected, significantly higher than for the vaccinated and unvaccinated cohorts. For subjects that survived their initial COVID-19 infection, the risk of testing positive for COVID-19 positive again was higher in the first 14-90 days for the post COVID-19 cohort compared with the vaccinated cohort. However, the hazard ratio for retesting COVID-19 positive between 90-300 days after the initial COVID-19 infection or after completing vaccination, for the post COVID-19 cohort was significantly lower than for the vaccinated cohort (HR 0.54, 95% CI 0.44-0.87, p<0.05). The unvaccinated cohort had, as expected, the highest risk of COVID-19 infection during this 300-day time period.

One goal of this study was to compare the risk of reinfection post COVID-19 infection with the risk of vaccine breakthrough infections. Vaccine breakthrough infection rates can serve as a risk benchmark, as these rates have been studied both in large prospective clinical trials and in ongoing observational studies. The findings of our analysis of community testing based reinfection and vaccine breakthrough rates can provide accurate insight for these relative rates of infection to the extent that the following hold true: that the proportion of undiagnosed infections is similar in both cohorts (which depends largely on equivalent willingness and access to testing in the two cohorts), that the rate of prior immunity from undiagnosed COVID-19 infection is similar in both cohorts, and that the risk of exposure to COVID-19 and specific variants was also similar in both cohorts. The goal of our matching cohorts based on date of infection/vaccination, age, gender, and postal code of residence was a means to minimize these potential areas of bias that would limit generalizability of our findings.

The matching of subjects based on date of initial COVID-19 infection or vaccination completion is of critical importance for limiting bias between the cohorts for risk of COVID-19 exposure and to different COVID-19 variants. This matching ensured that new positive tests were being recorded in all three cohorts during identical time periods of community-wide COVID-19 incidence and the presence of the same COVID-19 variants.

Matching for age was also of critical importance. Due to the age-based rollout of vaccination eligibility over time, the vaccinated group clearly has a strong deviation from the community demographics that changed over time. Our study design ensured we were comparing rates of death and COVID-19 test positivity for people of the same ages over identical time periods. For COVID-19 infection, age is probably the strongest predictor of mortality and matching on age is critical for this survival analysis. For post-testing COVID-19 positive, matching for age is likely also important for correcting differing exposure risks that may be age related (i.e., working vs. retired age groups), willingness to test, and access to healthcare of different age groups. Vaccination eligibility was also prioritized initially for persons with high-risk medical conditions. We were not able to correct for that factor with the data available. This health-status bias may explain the ongoing mortality in the vaccinated cohort that appears to be higher than in the post COVID-19 cohort.

Gender matching may also help to prevent biases for exposure risks and access to care. It is of interest that all three groups showed a female preponderance both prior to matching and after matching. While one could posit a higher risk of exposure for one gender or a biological predisposition for infection, that would not explain the female predominance for testing COVID-19 negative (Group 1), or for vaccination. This female bias may be due to willingness to access health care (vaccination and testing) or to a higher risk for non-COVID-19 conditions (such as seasonal allergies) that could lead to higher rates of COVID-19 testing. This female bias demonstrates the importance of our use of matching criteria.

Matching by postal zip code was used as a proxy for race/ethnicity and socioeconomic status, and also for proximity to health care services. There are significant differences between these demographic factors in Madera County by postal code area. We did not have income data available for subjects, and race/ethnicity data had too many incomplete entries to effectively match on this variable directly without excluding large numbers of persons. However, a post-matching comparison of the race and ethnicity data that is available shows that our matching process for the cohorts did create similar race and ethnicity demographic profiles, especially for the two cohorts of greatest interest with the most complete data for these variables (post COVID-19 infection and vaccinated group).

Our “unvaccinated” cohort is likely the least representative of a truly random sample of unvaccinated persons in Madera County. To have demographic information available and a defined cohort of people to observe for positive test events, we used persons who had completed a test prior to 1/1/2021 that was negative. It is likely that persons who have tested differ from the 31% of the county that had no test results recorded. Having tested negative (vs. never testing) suggests this group may have higher risk of exposure, better access to testing, and a greater willingness to test than the “never-tested” group. These biases could make this cohort have a higher incidence of positive COVID-19 test results than the group of all unvaccinated persons in Madera County.

The level of undiagnosed prior COVID-19 infections in each of our three cohorts cannot be assessed in this study. Prior COVID-19 infection would lead to greater than expected immunity in the unvaccinated and vaccinated cohorts. Prior COVID-19 infection is least likely to have occurred in our post COVID-19 cohort: as we demonstrate here a strong protection from reinfection in a post COVID-19 cohort, a prior undiagnosed COVID-19 infection would make a following diagnosed COVID-19 infection unlikely. Thus our finding of a hazard ratio of 0.54 post COVID-19 for retesting COVID-19 positive after 90 days compared with the vaccinated cohort would likely be strengthened, with an even lower HR, if our methods had allowed exclusion of prior undiagnosed COVID-19 infections from our three cohorts. This study did not evaluate the risks of COVID-19 infection after booster vaccination or after vaccination post COVID-19 infection. The results demonstrate that, post COVID-19, the risk of testing COVID-19 positive again is lower compared with vaccinated persons from 90-300 days post infection or vaccination. These results may not be applicable as new variants such as the Omicron variant emerge. These results do not contradict the strong recommendations for vaccination post COVID-19 infection, as other studies have demonstrated such vaccination leads to significantly greater protection from COVID-19 reinfection.

## Data Availability

All data produced in the present work are contained in the manuscript

